# Efficacy of propolis as an adjunct treatment for hospitalized COVID-19 patients: a randomized, controlled clinical trial

**DOI:** 10.1101/2021.01.08.20248932

**Authors:** Marcelo Augusto Duarte Silveira, David De Jong, Erica Batista dos Santos Galvão, Juliana Caldas Ribeiro, Thiago Cerqueira Silva, Andresa Aparecida Berretta, Thais Chaves Amorim, Raissa Lanna Araújo San Martin, Luis Filipe Miranda Rebelo da Conceição, Marcel Miranda Dantas Gomes, Maurício Brito Teixeira, Sergio Pinto de Souza, Marcele Helena Celestino Alves dos Santos, Márcio de Oliveira Silva, Monique Lírio, Lis Moreno, Julio Cezar Miranda Sampaio, Renata Mendonça, Silviana Salles Ultchak, Fabio Santos Amorim, João Gabriel Rosa, Paulo Benigno Pena Batista, Suzete Nascimento Farias da Guarda, Ana Verena Almeida Mendes, Rogerio da Hora Passos, for the BeeCovid Team

## Abstract

Among candidate treatment options for COVID-19, propolis, produced by honey bees from bioactive plant exudates, has shown potential against viral targets and has demonstrated immunoregulatory properties. We conducted a randomized, controlled, open-label, single center trial, with a standardized propolis product (EPP-AF) on hospitalized adult COVID-19 patients. Patients received standard care plus propolis at an oral dose of 400mg/day (n=40) or 800mg/day (n=42) for seven days, or standard care alone (n=42). Standard care included all necessary interventions, as determined by the attending physician. The primary end point was the time to clinical improvement defined as the length of hospital stay or oxygen therapy dependency. Secondary outcomes included acute kidney injury and need for intensive care or vasoactive drugs. Time in the hospital after intervention was significantly shortened in both propolis groups compared to the controls; median 7 days with 400mg/day and 6 days with 800mg/day, versus 12 days for standard care alone. Propolis did not significantly affect the need for oxygen supplementation. With the higher dose, significantly fewer patients developed acute kidney injury than in the controls (2 versus 10 of 42 patients). Propolis as an adjunct treatment was safe and reduced hospitalization time. The registration number for this clinical trial is: NCT04480593 (20/07/2020).

---

The coronavirus disease 2019 (COVID-19) pandemic continues to cause considerable morbidity and mortality. More than 75 million people have been infected with severe acute respiratory syndrome coronavirus 2 (SARS-CoV-2) globally, resulting in over 1.6 million deaths^1^ and unprecedented negative impacts on health care and the economy^2,3^. Despite advances in knowledge about viral targets for prospective medicines^4^, Covid-19 remains a considerable therapeutic challenge^5^. Critical features of this disease that have been investigated for medicinal intervention include viral spike protein interaction with cellular angiotensin-converting enzyme 2 (ACE2) and the human transmembrane protease TMPRSS2, which allow SARS-CoV-2 to attach to and enter host cells^6^. In later stages of COVID-19, an additional concern is a typical exaggerated inflammatory response, mediated by the “pathogenic” kinase PAK1^7^, which is associated with an increased need for intensive care and with high mortality rates^8^.

Propolis, a natural product made by bees from bioactive plant parts and resins, is already extensively consumed in various regions of the world, due to its reputation as a health aid,^7,9^ including purported antiviral activity^7,10^. Propolis components have shown promise for interfering with TMPRSS2 expression and ACE2 anchorage^9,11^, and they could help reduce inflammatory processes by PAK1 inhibition^7,12^. A pertinent objection to wider acceptance of propolis for medicinal use has been that it varies widely in composition and biological activity, according to the plant species that the bees selectively collect bioactive materials from in the different regions of the world^13,14^. In answer to this limitation, a standardized propolis product has been developed^15^ and tested in clinical trials^16,17,18^. Given the evidence concerning its biological properties, we hypothesized that propolis could help reduce the clinical impact of COVID-19 without interfering with other treatment options.

To evaluate the efficacy and safety of oral propolis for SARS-CoV-2 infection, we conducted a randomized, controlled, open-label trial in Brazil, Bee-Covid (The Use of Brazilian Green Propolis Extract (EPP-AF) in Patients Affected by COVID-19).

## Methods

### Trial Design and Oversight

Bee-Covid was a single-center, open-label, randomized, controlled trial conducted from June 3 through August 30, 2020, at São Rafael Hospital, Salvador, Bahia, in northeast Brazil. Because of the emergency nature of the trial, placebos were not prepared.

The protocol was approved by the Brazilian Committee of Ethics in Human Research (Registration number 31099320.6.0000.0048), and the trial was registered (ClinicalTrials.gov number, NCT04480593). The study was conducted in accordance with the principles of the Declaration of Helsinki and the Good Clinical Practice guidelines of the International Conference on Harmonization. All participating patients and/or legal representatives were informed about the objectives and risks of participation and gave written informed consent.

Eligible patients were randomly assigned in a 1:1:1 ratio to receive Standardized Brazilian Green Propolis Extract (EPP-AF®; Apis Flora Indl. Coml. Ltda, Ribeirão Preto, Brazil) for 7 days at 400 mg/day (one capsule of 100 mg, four times a day) plus standard care or 800 mg/day (two capsules of 100 mg, four times a day) plus standard care, or standard care alone (control group). The decisions on standard supportive treatment were made by the attending physicians, who were not involved in the study design or in the randomization process. Standard care comprised, as necessary, supplemental oxygen, noninvasive or invasive ventilation, corticosteroids, antibiotics and/or antiviral agents, vasopressor support, renal-replacement therapy, intra-aortic balloon pump and extracorporeal membrane oxygenation.

The dose of propolis was chosen based on studies that had used similar doses without observing adverse effects^16,17,19^. Patients were assessed daily during their hospitalization, from days 1 through 28. Data from patients who could not be reached for the 28-day follow-up were censored at hospital discharge. A standardized Brazilian green propolis extract, which is composed mainly of a green propolis produced in southeast Brazil, processed with a specific extraction and drying process, was selected for use in this study because of its batch-to-batch reproducibility^15^. The propolis components are characterized using high performance liquid chromatography (HPLC) to guarantee uniformity^15^.

### Randomization and masking

Randomization was stratified based on age, degree of pulmonary involvement, comorbidities, symptom onset time, and oxygen requirement as factors. The randomization scheme was generated by using the web site Randomization.com <http://www.randomization.com>. The randomization sequence was per block (using permuted blocks with four patients per block), including stratification. To minimize bias, allocation concealment was performed by researchers not involved in patient care. Maximum blindness among health professionals who had contact with the participants was sought; the professionals responsible for caring for patients did not have access to the intervention proposed in this study, and the data analysis was carried out with external statistical support and in an impartial manner.

### Patients

Hospitalized patients over 18 years of age diagnosed with SARS-CoV-2 infection, confirmed by polymerase chain reaction-reverse transcriptase testing, were considered eligible if symptoms started within 14 days of the randomization date. Exclusion criteria included pregnancy or lactation, known hypersensitivity to propolis, active cancer, human immunodeficiency virus carriers, patients undergoing transplantation of solid organs or bone marrow or who were using immunosuppressive medications, bacterial infection at randomization, sepsis or septic shock related to bacterial infection at randomization, impossibility of using the medication orally or by nasoenteral tube, known hepatic failure or advanced heart failure (New York Heart Association [NYHA] class III or IV).

### Outcome Measures

The primary end point was the time to clinical improvement defined as the length of hospital stay or oxygen therapy dependency time. Secondary end points were the percentage of participants requiring mechanical ventilation, rate of acute kidney injury, need for renal replacement therapy, need for intensive care treatment, and need for vasoactive drugs. We also analyzed laboratory parameters, including variation in serum levels of C-reactive protein over the seven days after randomization (see Supplementary Table S2 and Fig. S2), and death.

Acute kidney injury was defined according to Kidney Disease: Improving Global Outcomes (KDIGO) criteria as stage 1 (increase in serum creatinine by 0.3 mg/dl within 48 hours or a 1.5 to 1.9 times increase in serum creatinine from baseline within 7 days), stage 2 (2.9 times increase in serum creatinine within 7 days), or stage 3 (3 times or more increase in serum creatinine within seven days or initiation of renal replacement therapy)^20^. Safety outcomes included adverse events that occurred during treatment, serious adverse events, and premature or temporary discontinuation of treatment. Adverse events were classified according to the National Cancer Institute Common Terminology Criteria for Adverse Events, version 4.03.

### Statistical Analysis

As information about the use of propolis for respiratory syndrome conditions was limited, we used data from previous studies to infer the length of hospital stay due to COVID-19^21^. We assumed a mean (± standard deviation) length of hospital stay of 13 ± 6.5 days in the control group. Based on a two-sided type I error of 0.05 and 80% power to identify a difference of four days of length of hospital stay between the lower dose and the control groups, a sample size of 42 patients by group would be needed. This number of patients were recruited for the group with the higher propolis dose.

The primary analysis study population comprised all patients who had been randomized (intention-to-treat population), using the group to which a patient was allocated as a variable, regardless of the medication administered or treatment adhesion. The main objective was to evaluate the effectiveness of propolis in reducing the number days of oxygen therapy and length of hospital stay in adult patients with confirmed SARS-CoV2 infection. The number of days on oxygen therapy was based on the number of days patients were on invasive or noninvasive oxygen therapy, both counted after randomization. The time to discharge or no further need for supplemental oxygen was assessed after all patients had reached day 28. Patients still dependent on oxygen therapy or still hospitalized or who died by the end of follow-up time were considered as time equal 28 days in the analysis. The endpoints were assessed visually using Kaplan Meier curves. The treatment effect was presented as the mean difference, with 95% confidence intervals and p-values. We used a generalized linear model with a Gamma distribution, considering age and the treatment groups as independent variables.

The binary outcomes were assessed with a logistic-regression model. Continuous outcomes were evaluated through linear regression. Ventilator time, intensive care unit time, and vasopressor drug use time were adjusted additionally for the status at randomization. Adverse events were expressed as counts and percentages and compared between groups using the Fisher exact test. Analyses were performed with R software, version 4.0.2 (R Project for Statistical Computing).

## Results

### Patients

Of the 242 patients who were assessed for eligibility, 125 were enrolled and underwent randomization and 124 began treatment; 40 patients were assigned to receive the lower dose of propolis (400 mg/day), 42 to receive the higher dose of propolis (800 mg/day) and 42 to receive standard care alone (control group) (Fig. 1). One patient was excluded after randomization and before receiving the medication (withdrew consent after randomization). The mean (±standard deviation) age of patients in this trial was 50.0 ± 12.8 years, and 69.4% of the patients were men (Table 1). Overall, 5.2% of patients had hypertension, 51.6% were obese, 21.0% had diabetes and 7.3% had chronic pulmonary obstructive disease. The median (interquartile range) time from symptom onset to randomization was 8 (6 to 10) days. At randomization, 3.2% were using invasive mechanical ventilation, 48.4% were receiving oxygen with non-invasive ventilation and 41.1% were being treated in the intensive care unit.

**Table 1.** Demographic and clinical characteristics of the COVID-19 patients at baseline. Abbreviations: BMI, body-mass index (calculated as weight in kilograms divided by height in meters squared), COPD, chronic obstructive pulmonary disease, CT computerized tomography, SpO2, Peripheral oxygen saturation, ^a^lopinavir-ritonavir, remdesivir, tocilizumab, and colchicine were not used for any patient.

| **Variables** | **Total**<br>**(N=124)** | **Standard care**<br>**(N=42)** | **EPP-AF**<br>**400mg/day (N=40)** | **EPP-AF**<br>**800mg/day (N=42)** |
| --- | --- | --- | --- | --- |
| **Age, mean (SD), y** | 50.0 (12.8) | 51.6 (14.3) | 49.5 (12.8) | 48.9 (11.2) |
| **Male sex, No. (%)** | 86 (69.4) | 28 (66.7) | 28 (70.0) | 30 (71.4) |
| **Ethnicity, No. (%)** |  |  |  |  |
| White | 33 (30.6) | 10 (23.8) | 13 (32.5) | 10 (23.8) |
| Black | 26 (21.0) | 12 (28.6) | 5 (12.5) | 9 (21.4) |
| Mixed | 65 (52.4) | 20 (47.6) | 22 (55.0) | 23 (54.8) |
| **Coexisting conditions, No. (%)** |  |  |  |  |
| Diabetes | 26 (21.0) | 11 (26.2) | 10 (25.0) | 5 (11.9) |
| Hypertension | 56 (45.2) | 21 (50.0) | 18 (45.0) | 17 (40.5) |
| COPD/Asthma | 9 (7.3) | 2 (4.8) | 3 (7.5) | 4 (9.5) |
| Obesity | 64 (51.6) | 18 (42.9) | 23 (57.5) | 23 (54.8) |
| **Median time from symptom**<br>**onset to randomization, median**<br>**(IQR)** | 8.0 (6.0-10.0) | 8.0 (6.0-10.7) | 8.0 (5.0-9.0) | 9 (7.0-10.0) |
| **Randomization location, No. (%)** |  |  |  |  |
| Ward | 73 (58.9) | 22 (52.4) | 27 (67.5) | 24 (57.1) |
| ICU | 51 (41.1) | 20 (47.6) | 13 (32.5) | 18 (42.9) |
| **Conditions at randomization,**<br>**No. (%)** |  |  |  |  |
| No additional oxygen therapy | 60(48.4) | 20 (47.6) | 20(50.0) | 20(47.6) |
| Nasal cannula | 59(47.6) | 21(50.0) | 17 (42.5) | 21 (50.0) |
| High-flow nasal cannula | 1 (0.8) | 0 (0.0) | 1 (2.5) | 0 (0.0) |
| Invasive ventilation | 4 (3.2) | 1 (2.4) | 2 (5.0) | 1 (2.4) |
| **Temperature, median (IQR), °C** | 36.3 (35.7-36.8) | 36.4 (35.9-36.8) | 36.2 (35.7-36.8) | 36.2 (35.5-37.0) |
| **Respiratory rate >24/min — No.**<br>**(%)** | 12 (9.7) | 5 (11.9) | 4 (10.0) | 3 (7.1) |
| **SpO2 < 93%, No. (%),** | 21 (16.9) | 4 (9.5) | 6 (15.0) | 11 (26.2) |

**Table 1.** Demographic and clinical characteristics of the COVID-19 patients at baseline.
| Variables | Total<br>(N=124) | Standard Care<br>(N=42) | EPP-AF 400mg/day<br>(N=40) | EPP-AF<br>800mg/day (N=42) |
| --- | --- | --- | --- | --- |
| **Lung parenchyma involvement**<br>**estimated by CT, No. (%)** |  |  |  |  |
| < 25% | 29 (23.4) | 10 (23.8) | 13 (32.5) | 6 (14.3) |
| 25-50% | 62 (50.0) | 20 (47.6) | 14 (35.0) | 28 (66.7) |
| 50-75% | 29 (23.4) | 11 (26.2) | 10 (25.0) | 8 (19.0) |
| >75% | 4 (3.2) | 1 (2.4) | 3 (7.5) | 0 (0.0) |
| **Concomitant COVID-19**<br>**treatment<sup>a</sup>, No. (%)** |  |  |  |  |
| Azithromycin | 118 (95.2) | 41 (97.6) | 37 (92.5) | 40 (95.2) |
| Chloroquine or<br>Hydroxychloroquine | 4 (3.2) | 2 (4.8) | 0 (0.0) | 2 (4.8) |
| Oseltamivir | 76 (61.3) | 28 (66.7) | 24 (60.0) | 24 (57.1) |
| Corticosteroids | 100 (80.6) | 39 (92.9) | 26 (65.0) | 35 (83.3) |
| **Creatinine (mg/dl), median**<br>**(IQR)** | 0.81 (0.62-1.00) | 0.68 (0.55-1.02) | 0.79 (0.67-1.00) | 0.85 (0.76-1.03) |
| **White-cell count (x10<sup>3</sup>/mm<sup>3</sup>),**<br>**median (IQR)** | 5.9 (4.3-7.9) | 6.2 (4.4-7.8) | 5.3 (3.6-7.1) | 6.1 (4.8-8.2) |
| **Lymphocyte count (x 10<sup>3</sup>/ mm<sup>3</sup>),**<br>**median (IQR),** | 0.9 (0.7-1.3) | 0.82 (0.7-1.0) | 1.0 (0.7-1.4) | 1.0 (0.7-1.3) |
| **Platelet count (x 10<sup>3</sup>/ mm<sup>3</sup>),**<br>**median (IQR)** | 183.0 (142.5-233.5) | 164.0 (131.0-224.2) | 201.5 (145.7-239.5) | 188.5 (154.2-235.7) |
| **Aspartate aminotransferase**<br>**(U/L) — median (IQR)** | 43.0 (32.5-63.0) | 46.5 (31.7-65.2) | 49.0 (31.5-73.5) | 39.0 (33.0-49.5) |

**Figure 1.**
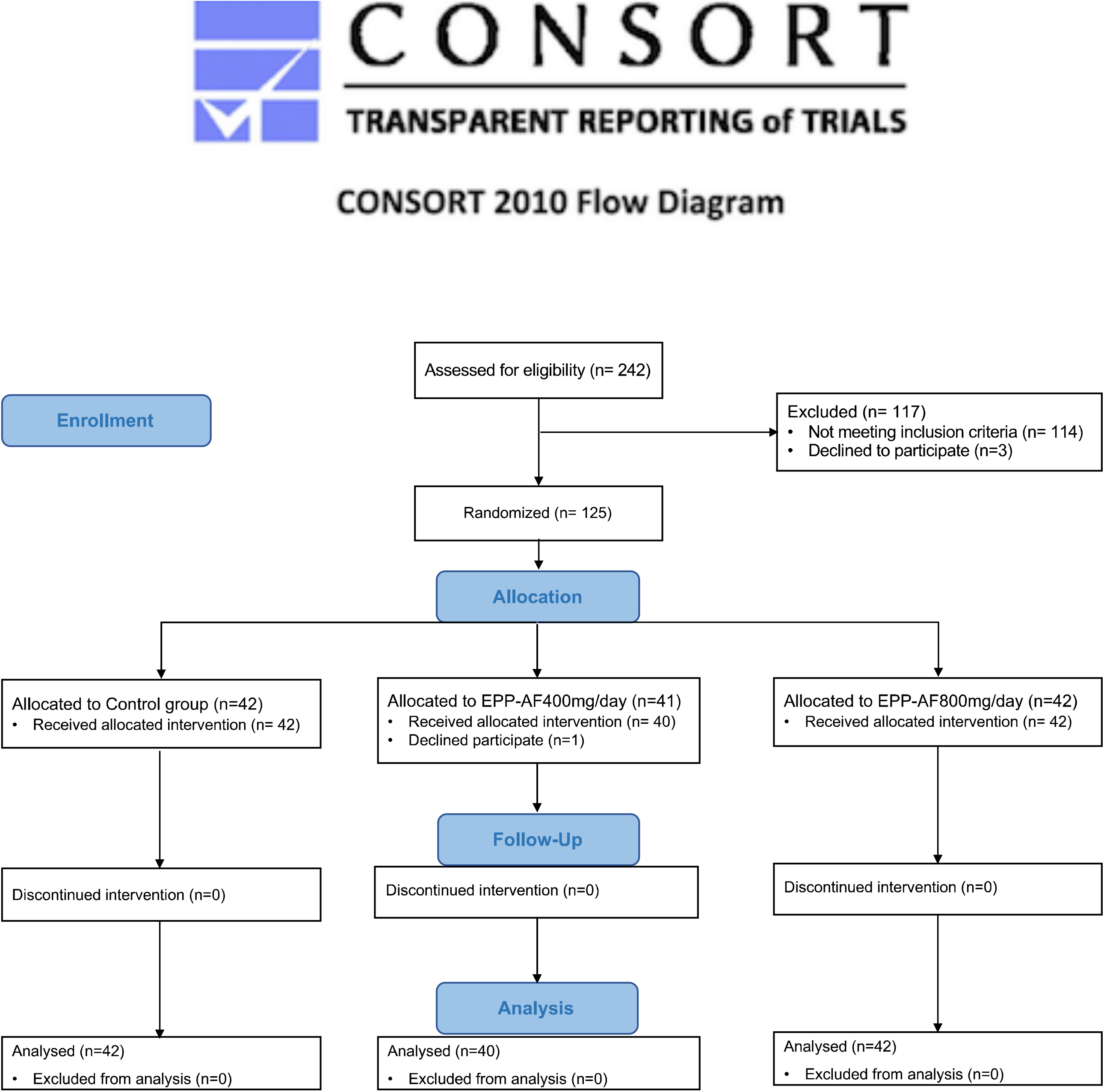
Enrollment and randomization of COVID-19 patients.

Follow-up information at day 28 after admission for the primary outcome was complete for all 124 patients. The use of azithromycin, chloroquine or hydroxychloroquine and oseltamivir was similar in all groups. The frequency of patients requiring use of corticosteroids was lower in the group receiving the lower dose of propolis (65%) when compared to the higher dose of propolis (83%) and standard care (93%) groups.

### Primary outcomes

Length of hospital stay at 28 days was significantly shorter in both groups receiving propolis than in the standard care group, with a mean difference between the lower propolis dose and the control of −3.03 days (95% confidence interval [CI] −6.23 to −0.07; p=0.049; median 7 versus 12 days), and for the higher dose of propolis compared to control it was - 3.88 days (95% CI −7.00 to −1.09; p=0.009; median 6 versus 12 days,) (Table 2). The cumulative frequency of discharge from the hospital is shown in Fig. 2a.

**Table 2.**
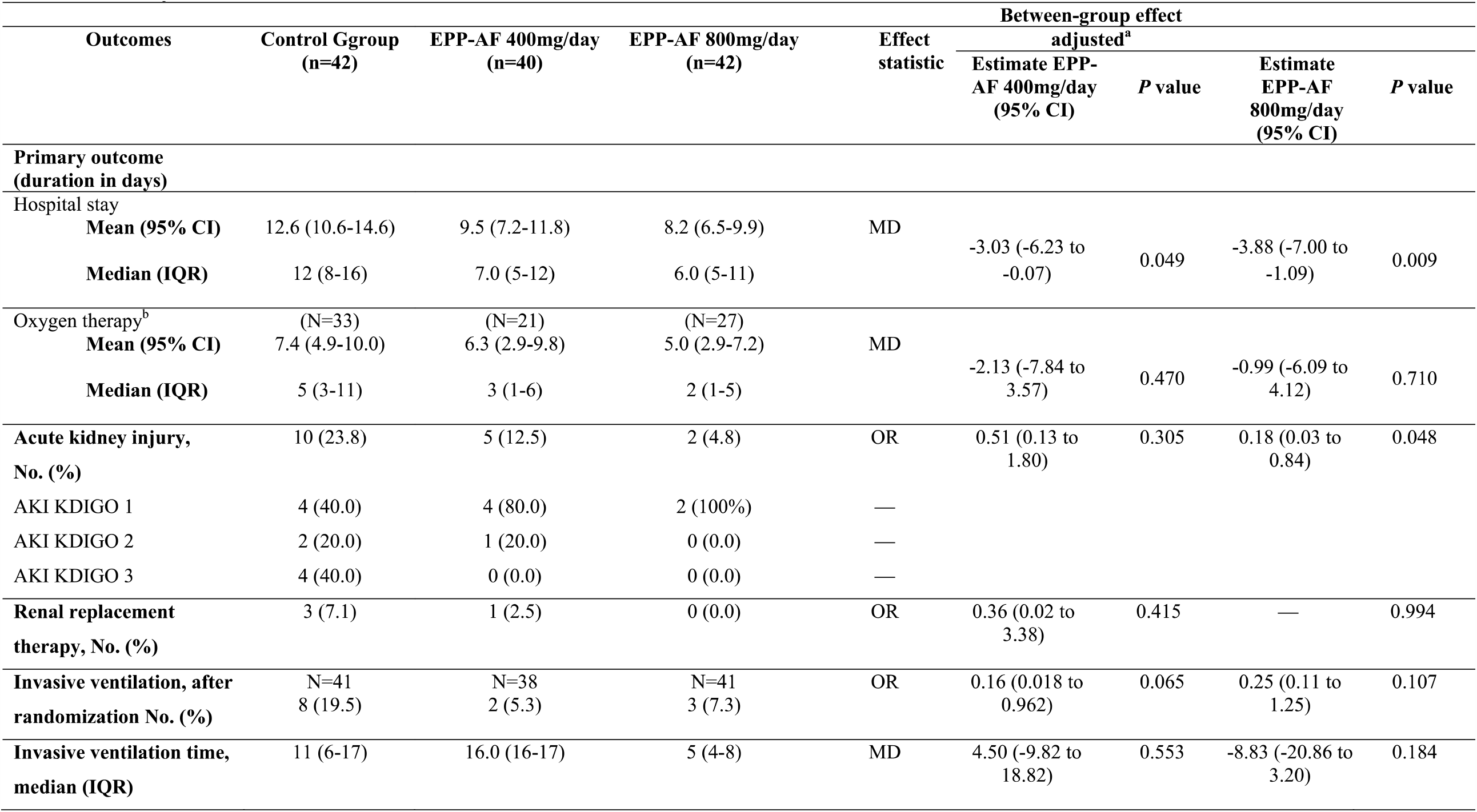
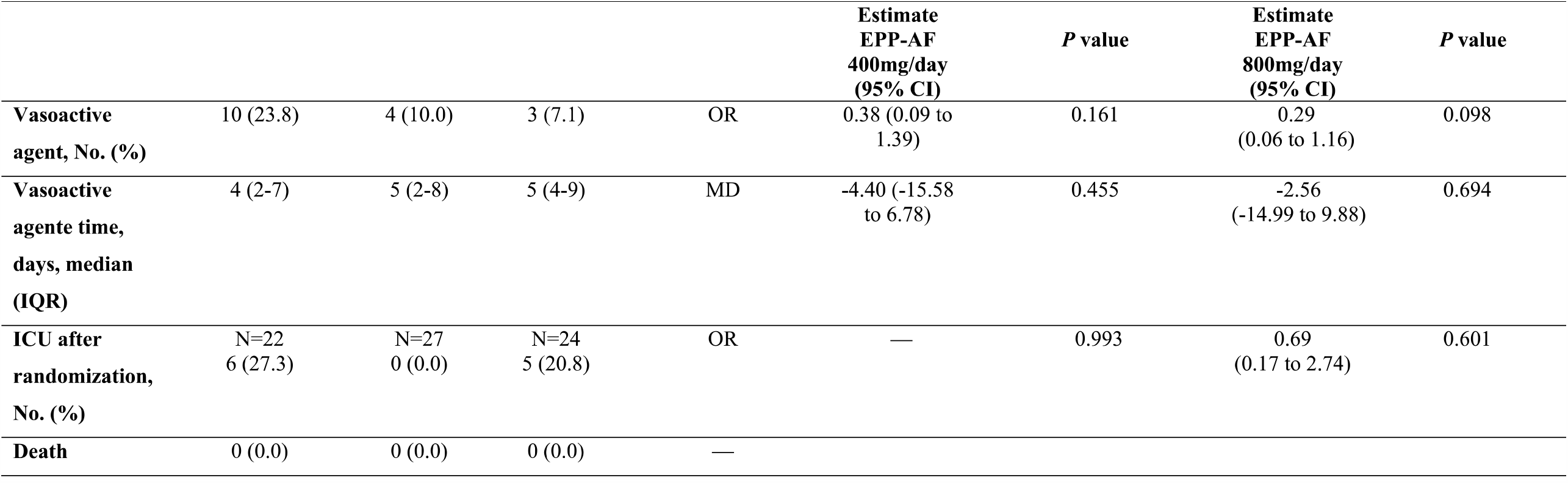
Study outcomes MD = mean difference; OR = odds ratio. ^a^ All models were adjusted for age. ^b^For invasive ventilation time and vasoactive agent time, the models are adjusted for age and randomization location. Seven inpatients 28 days after admission | Four patients on oxygen dependence 28 days after admission.

**Figure 2.**
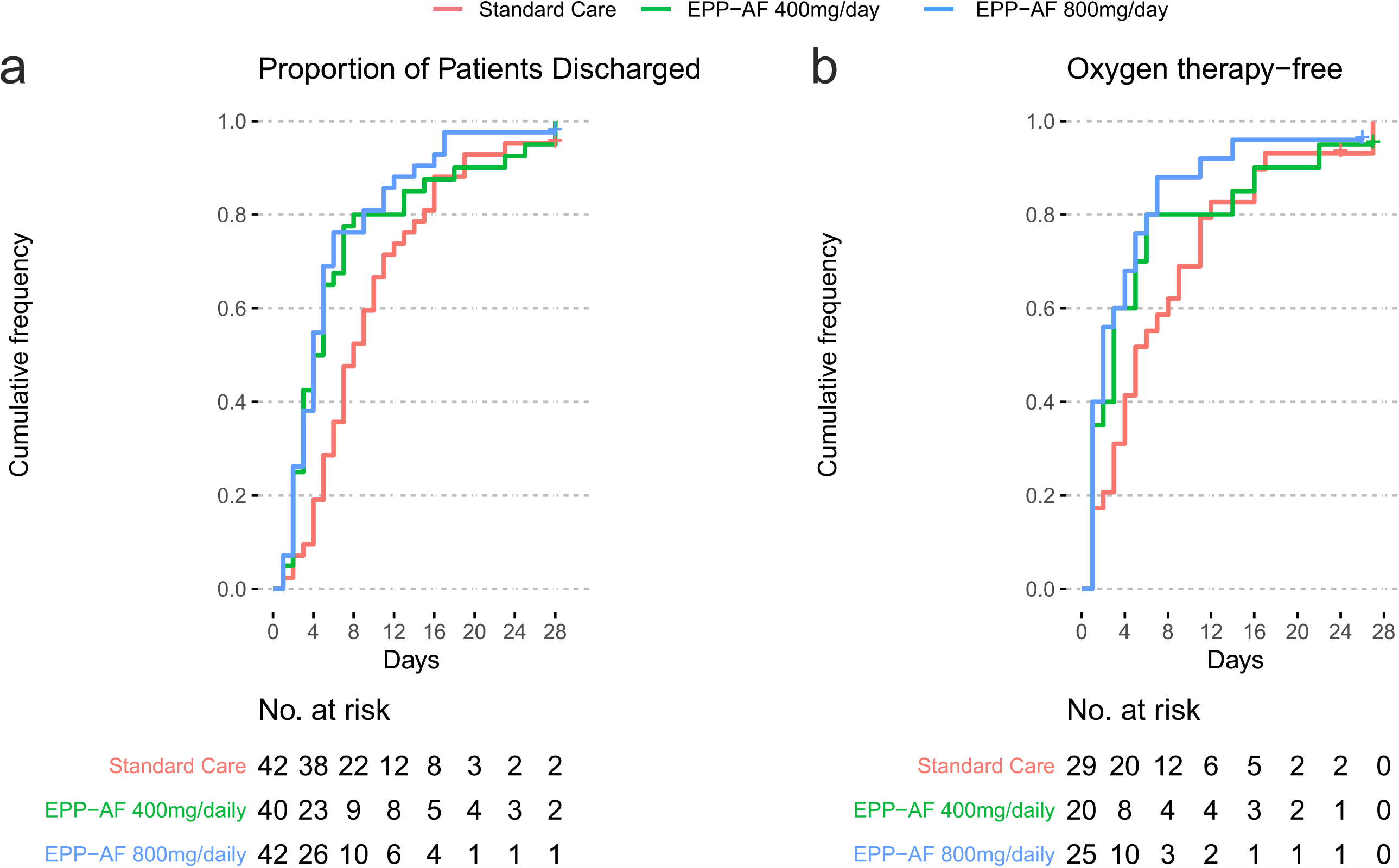
Primary outcomes for the COVID-19 patients: Length of hospital stay at 28 days (Figure 2a) and oxygen therapy dependency time (Figure 2b).

Patients assigned to propolis did not have a significantly different time on oxygen (with or without invasive ventilation) compared to the control group. The mean difference for the lower dose of propolis versus control was -2 days (95% CI −7.84 to 3.57; p=0.470), and for the higher dose of propolis versus control it was - 0.99 days (95% CI −6.09 to 4.12; p=0.710) (Table 2). The cumulative frequency of patients no longer on oxygen is shown in Figure 2b.

### Secondary outcomes

The incidence of acute kidney injury was 23.8, 12.5, and 4.8% for the control, 400 and 800 mg propolis/day groups, respectively. Only the higher propolis dose group had a significantly lower rate of acute kidney injury than the control group (odds ratio [OR] 0.18; 95% CI 0.03 to 0.84; p=0.048).

There were no significant differences in any of the remaining secondary outcomes. None of the patients taking the lower dose of propolis needed to be transferred to the intensive care unit, while the rate was 20.8% in the higher propolis dose group compared to 27.3% in the control (OR 0.69; 95% CI 0.17 to 2.74; p=0.601) (Table 2). A total of 13 patients initiated mechanical ventilation after randomization (5.3% of patients assigned the lower dose of propolis, 7.3% using the higher dose of propolis and 19.5% in the control group). The median number of days of invasive ventilation after randomization was 16 (16 to 17) in the group assigned to propolis 400mg/day, 5 (4 to 8) in the propolis 800 mg/day and 11 (6 to 17) in the control group (Table 2). The need for vasoactive agents was numerically lower in the groups receiving propolis than in the control group (10% with propolis 400mg/day; 7.1% with propolis 800mg/day and 23.8% in the control). The time of vasoactive drug use and intensive care unit necessity was similar in all groups (Table 2). Laboratory data, including variation in serum levels of C-reactive protein over the seven days after randomization was recorded (see Supplementary Table S2 and Fig. S2).

### Safety outcomes

Adherence to the trial intervention did not differ according to the treatment group. No patient had propolis treatment discontinued due to side effects. The percentages of patients experiencing adverse events did not differ significantly among the three groups. The most severe adverse event overall was shock/ use of vasoactive drugs in 23.8% of the patients in the standard care group versus 10% in the propolis 400 mg/day group and 7.1% in the propolis 800 mg/day group; p = 0.098. The second most common adverse event was acute respiratory failure, which occurred at a rate of 19.5% in the standard care group, 5.3% with propolis 400 mg/day and 7.3% with propolis 800 mg/day (Table 2).

Gastrointestinal adverse events, specifically nausea, presented in one patient in the group with the lower dose of propolis and one patient in the control group. The only neurologic event was headache, and it presented in only one patient in the control group. The percentages of patients with laboratory abnormalities were similar in the three groups (Supplementary Table SI and Fig. SI). Episodes of itching, an increase in alkaline phosphatase, and rash were not observed in any of the patients. No patient had propolis treatment discontinued due to any adverse event.

## Discussion

Through this randomized clinical trial, we found that oral administration of propolis for 7 days was safe and beneficial. Propolis plus standard support was associated with a reduction in length of hospital stay after randomization for treatment, median 7 days (5 to 12) with 400 mg/day and 6 days (5 to 11) with 800 mg/day, compared with a median of 12 days (8 to 16) for standard treatment alone.

Propolis is a resinous substance produced by bees from plant parts and resins that they use to protect the hive. It has antiviral, anti-inflammatory, immunoregulating, antiproteinuric and antioxidant properties^7,17,18,22^. Normally, propolis varies according to climate regions and to the types of plants available^9,19^. Differences between propolis products, due to a lack of standardization involving the botanical source, solvent extraction and processing methods was a challenge identified by the European Medicine Agency, since it would be difficult to extrapolate the available safety and efficacy information for all types of propolis^9^. In order to overcome this problem with the variability of propolis, a standardized propolis product that is chemically and biologically reproducible was developed^15^; it has proven efficacy and minimal interaction with medications in clinical studies^16,17,19^.

Severe pulmonary involvement is the most common problem associated with advanced cases of COVID-19^23,24^. All 124 patients included in our study had some degree of pulmonary involvement, and just over half of them were on oxygen support at randomization, demonstrating that this patient population had moderate to severe cases of this disease. The time under oxygen support, including invasive and non-invasive therapy was not significantly different between the groups; the median in the 400mg/day group was three days (1 to 6), and in the 800mg/day group it was two days (1 to 5) (p= 0.470 and 0.710, respectively), compared with five days (3 to 11) for standard care alone. There was an apparent tendency for patients treated with propolis to have a reduced need for invasive oxygen therapy; but since relatively few patients required this type of support overall, we cannot conclude that propolis was beneficial based on this clinical parameter.

Acute kidney injury is a common complication in COVID-19^8,25^. Its incidence varies among COVID-19 patients, being associated with a poor prognosis, longer hospitalization times and greater mortality^8,23,26^. An observational study of more than 5,000 hospitalized COVID-19 patients reported an overall frequency of 36% acute kidney injury; among patients on noninvasive oxygen support, the rate was 20%, and among those on mechanical ventilator support it was 89%. This implies a “cross talk” between lung and kidney under inflammatory insult^26,27^.

Patients treated with the higher dose of propolis had a significantly lower incidence of acute kidney injury compared to the control group. These findings have an important clinical significance, since acute kidney injury is associated with the worst outcomes^25,26^. The development of severe kidney lesions in COVID-19 patients is multifactorial, involving risk factors inherent to these patients (e.g., comorbidities), including volemic status (circulating blood volume), exposure to nephrotoxins, acute cardiac involvement (cardiorenal syndrome), systemic inflammation (immune response dysregulation; cytokine storm), endothelial lesions (microthrombi formation), and renal tubular lesions^26,28^.

There is an urgent need to find treatment options for COVID-19 that are safe and potentially able to interfere with viral targets and reduce viral virulence and damage to the body. ACE2 and TMPRSS2 are essential for invasion of SARS-CoV-2 into host cells. TMPRSS2 cleaves the spike protein (S) of SARS-CoV-2, which facilitates the fusion of SARS-CoV-2 with the cell membranes and entry into the cell via ACE2^6,29^. Within cells, the coronavirus promotes activation of PAK1, a kinase that mediates the inflammatory response, pulmonary fibrosis and other critical mortality factors. Higher levels of PAK1 also reduce the adaptive immune response, facilitating viral replication^8^. Various propolis components can inhibit and/or modulate these viral targets^7,30^.

Our study has several limitations. It was a single center clinical trial, requiring greater caution in interpretations and generalizations concerning the findings. Although we blinded most of the health professionals involved in care of the patients, to reduce the possibility of interference, this trial was open. Also, the patients were followed for only a short period, limiting the possibility of evaluating long-term effects.

In conclusion, the addition of oral propolis to the standard care procedures was safe and had clinical benefits for the hospitalized COVID-19 patients, especially evidenced by a reduction in the hospitalization time. Possibly, administration early in the disease course would have an even greater effect in reducing the disease’s impact. Given our findings, and the evidence concerning the ways in which propolis can affect various disease mechanisms that are relevant to SARS-CoV-2 infection, propolis should be considered as an adjuvant in the treatment of COVID-19 patients. Future studies to further assess the impact of propolis on renal protection would be useful.

## Data Availability

Relevant data is available in the manuscript file. If additional data is needed, it can be obtained from the Corresponding Author: Marcelo Augusto Duarte Silveira, MD, PhD, D'Or Institute for Research and Education (IDOR), Hospital Sao Rafael, Sao Rafael Avenue, 2152 - Sao Marcos, Salvador - Bahia, 41253-190, Brazil

## Acknowledgments

The authors thank the patients for their valuable contributions. The authors also thank Apis Flora Indl. Coml. Ltda. for funding and for providing the propolis, and the D’Or Institute for Research and Education (IDOR) and Hospital São Rafael for funding and their staff for assistance.

## Author contributions

M.A.D.S. designed the trial and was the principal investigator, with overall responsibility for conducting the trial and for medical oversight of trial implementation. M.A.D.S., R.H.P., S.N.F.G., A.V.A.M., J.G.R., and P.B.P.B. were responsible for the study design. M.A.D.S., D.D.J., A.A.B. and R.H.P. wrote the final report. J.C.R. and T.C.S. were responsible for analyzing the data. E.B.S.G., R.L.A.S.M., and T.C.A. were responsible for coordinating and organizing the participants’ data. S.P.S., L.F.M.R.C., M.M.D.G., M.B.T., M.H.C.A.S., M.O.S., M.L., S.S.U., and F.S.A. contributed to the clinical and operational implementation of the trial. All authors contributed to trial design and interpretation of data and reviewed the final report.

